# Mechanistic view on the influence of fluctuations in outdoor temperature on the worsening of the course of the disease and hospitalizations associated with the SARS-CoV-2 Omicron wave in 2022 in the Tomsk region, Russia

**DOI:** 10.1101/2023.01.04.23284173

**Authors:** A.N. Ishmatov, A.A. Bart, S.V. Yakovlev

## Abstract

It is well known that low air temperatures negatively affect the human respiratory system and can suppress protective mechanisms in airways epithelial cells.

In this study, we put forward the hypothesis that the ‘compromised airway epithelium’ of infected persons can be extremely sensitive to external influences and therefore can be used as an “indicator” and serve to investigate the impact of low air temperatures (as and other external factors) on the respiratory system.

Here we supposed that a short-term impact of drop in outdoor temperature on the ‘compromised airway epithelium’ should lead to increased symptoms and severity of the disease.

We have analyzed a short-term impact of the air temperature drop on the worsening of disease in patients with COVID-19 (indicated by bursts of daily hospitalizations), which fell on the main epidemic wave in 2022 associated with SARS-CoV-2 Omicron variant.

It was found that even a small and/or short-term impact of drop in outdoor daily temperatures can lead to increased symptoms and severity of the disease (COVID-19).

We have identified 14 characteristic points (days) where the temperature drop was more than 3 degrees during the main pandemic wave in 2022. It was shown that each characteristic points clearly associated with characteristic bursts in the number of daily hospitalizations with a time lag of 1-2 days.

Thus, it was found that the results of the study can be used in predicting a sudden increase in the number of hospitalizations, which can be used to timely warn clinics and medical hospitals for an increase in the number of seriously ill patients.

The findings can be used to improve systems to prevent additional risks connected with impact of drop in air temperature on worsening disease in patients and infected people who do not have or have mild or subtle symptoms of the disease – especially during an epidemic or pandemic wave.

## Introduction

Novel coronavirus disease (COVID-19) is a respiratory infection caused by severe acute respiratory syndrome coronavirus 2 (SARS-CoV-2). COVID-19 is a complex disease with clinical responses varying from asymptomatic to severe with catastrophic respiratory failure and death (Bridges et al., 2022). The COVID-19 pandemic has resulted in an extraordinary incidence of morbidity and mortality, with 645 million confirmed cases and 6.6 million deaths worldwide (https://covid19.who.int/).

During the natural evolution of the COVID-19 pathogen, dominant variants emerge that account for most new infections. The Omicron variant has become the dominant agent of the wave of the COVID-19 pandemic in the beginning of 2022 (Del Rio & Malani, 2022).

It is well known that meteorological factors may influence the incidence of many infectious diseases. It has been suggested that the weather conditions has contributed to spreading the SARS-CoV-2 as a result of the same drivers that contribute to the occurrence of respiratory viruses, including seasonal influenza and coronaviruses (see related reviews in (Xie & Zhu, 2020; Christophi et al., 2021; Kaplin et al., 2021; Nichols et al., 2021)).

It was assumed that the dynamics of the COVID-19 pandemic suggests that a high percentage of susceptible people at the beginning of the pandemic means that the effect of the weather was relatively small, but this effect increases in next years as the SARS-CoV-2 becomes endemic and the population’s susceptibility to the virus declines (Baker et al., 2020; Nichols et al., 2021).

The seasonality of respiratory viruses and influenza has been extensively researched and continues to be investigated. Specifically, ambient temperature is considered to be the key climate factor that affects seasonality of respiratory infections. Nevertheless, possibly due to the differences in latitudes and climate regions, the relationship between ambient temperature and respiratory infections was not consistent. Seasonality of respiratory viruses commonly connected with cold seasons in the Northern Hemisphere and wet seasons in tropical countries – these findings provide a simple climate-based model rooted in empirical data that accounts for the diversity of seasonal influenza patterns: “cold-dry” and “humid-rainy” (see related reviews in (Eccles, 2002b; Lofgren et al., 2007; Lipsitch and Viboud, 2009; Tamerius et al., 2013; Moriyama et al., 2020; Qi et al., 2022; Li et al., 2022)).

The seasonality of seasonal coronaviruses has been examined too in many countries across the world and showed seasonal patterns similar to influenza and respiratory syncytial virus (RSV) in temperate climates (see review in (Li et al., 2020; Nichols et al., 2021)). The study by Nichols et al. (2021) has found that seasonal coronavirus infections in England and Wales have a broadly similar distribution to influenza A and human bocavirus infections. It has been suggested that forecasting based on weather and climatic conditions (global radiation, relative and/or absolute humidity, air temperature, sunshine (hours per day), dewpoint temperature, precipitation and etc.) might help in preparing health care systems and populations to better protect those most at risk from serious complications from becoming infected (Scafetta, 2020; Nichols et al., 2021).

It is important to note that while the many studies show associations between some weather parameters and respiratory infections (including COVID-19) it is recognized that the drivers of infection and seasonal epidemics involve combined effects of physical, physiological, social and behavioral elements (Bontempi, 2020; Ishmatov, 2020a; Bontempi and Coccia, 2021; Culqui et al., 2022). Moreover, many of the weather factors auto-correlate to some extent because of their natural relationships in the environment – for example, relative humidity is influenced by temperature; temperature can be influenced by sunshine and etc. (Nichols et al., 2021).

Despite a lot of research in this area the seasonality (predictable fluctuation or pattern that recurs or repeats over a one-year period) of COVID-19 is still poorly understood and insufficiently taken into consideration (D’Amico et al., 2022).

In this study, we put forward the hypothesis of the ‘compromised airway epithelium’ and the impact of outdoor temperature as the last straw in the disease course.

The hypothesis is as follows the airway epithelium of an infected person at the first stages of the infection/disease and in the mild and moderate course of the disease (when there are no symptoms or they are insignificant) partially loses its functionality. In such a state, the balance of the functioning of the airway epithelium (as well as its defense mechanisms) can be easily disturbed by different external triggers. This can lead to an even greater weakening of the protective functions in the airway epithelium and, as a result, to the uncontrolled development of the infection.

Thus, here we suppose that even a small and/or short-term impact of drop in outdoor temperature (as well as other factors) on the ‘compromised airway epithelium’ should lead to increased symptoms and severity of the disease.

**The aim of the study is to find the relationship between the influence of outdoor temperature (as one of the weather factors) and the increased symptoms and the deterioration of the course of diseases during pandemic wave associated with the SARS-CoV-2 Omicron strain in 2022**

Here we based on the mechanistical point of view on how the air temperature could affect the respiratory system and lead to the worsening of the course of the disease and hospitalizations associated with the COVID-19. It is well known that that the respiratory epithelial cells can be critically cooled by inhaled cold/cool air and it can lead to the reduction of antiviral response in the cells, the inhibition of mucociliary clearance (Tyrrell and Parsons, 1960; Salah et al., 1988; Eccles, 2002b; Mourtzoukou and Falagas, 2007; Makinen et al., 2009; Foxman et al., 2015; 2016; Iwasaki et al., 2017). Foxman et al. (Iwasaki lab) (Foxman et al., 2015; Foxman et al. 2016) (Iwasaki et al., 2017) had clearly shown the mechanism of reducing the immune response of cells in the respiratory tract of mice during cooling the cells. The ability of various strains of rhinoviruses replicate more better in the respiratory epithelial cells at 33 °C than at the normal lung temperature of 37 °C. It is also important to note here the mechanism of influence of personal and outdoor temperature exposure during cold and warm seasons on lung function and respiratory symptoms in COPD which was described by Scheerens et al. (2022).

## Methods

We limited our analyses to the city of Tomsk and the Tomsk agglomeration. Tomsk (56°30′N 84°58′E) is a city and the administrative center of Tomsk Oblast in Russia, located on the Tom River. The city’s population is 570776 at the end of 2021, with a population density of 1937 person/km2. Tomsk has a humid continental climate barely escaping a subarctic classification. The annual average temperature is +1.2 °C. The total annual rainfall is 587 millimeters (URL:https://en.wikipedia.org/wiki/Tomsk (date of access: Jun 1, 2022)).

We considered the pandemic period from January 1st, 2022, to March 31st, 2022, associated with the SARS-CoV-2 Omicron strain, the dominant agent of the new wave of the COVID-19 pandemic in 2022 in the region.

The data on measured outdoor temperatures were collected from official reports on weather status are issued by Regional Directorates of Hydrometeorological Service in Russia: http://www.meteorf.ru/about/structure/local/.

To analyze daily dynamics in the symptom aggravation and disease severity, we used data on daily hospitalizations associated with COVID-19. The statistical data on COVID-19 hospitalizations were obtained from open official sources for Tomsk and the Tomsk region (URL: https://gogov.ru/covid-hosp-stats/tms#data (In Russian - date of access: Jun 1, 2022)). Here we hypothesized that short-term fluctuations in outdoor temperature may be associated with an increase in the symptoms of patients with COVID-19 and are associated with the presence of pneumonia at the time of diagnosis of COVID-19 in hospitals (for those cases that fall into official statistics). In our work, we hypothesized that many COVID-19 pneumonia diagnoses are triggered by activity in the preceding hours.

Using a control group of patients for the analysis is a difficult task. And we wondered what would be the best representative of the population that set the precedents? − The simple answer was the cases themselves. The case-crossover study design is an adaptation of the case-control study, in which cases serve as their own controls, and is well suited to the study of transient risk factors. We used a parameter of hospitalization and pneumonia as a proxy for the severity of COVID-19 symptoms. Although this may be justified on the grounds that the most severe forms of COVID-19 usually lead to pulmonary manifestations on their own (we did not have any direct measures of severity or a confirmed severity index). We have adapted this approach; this is the same approach that has been proposed for the analysis of cases of heart attacks and severe illness by Maclure (1991), this method was also used in a recent study on individual emergency department visits for COVID-19 (Lavigne, et al., 2022).

## Results and Discussion

A graph of outdoor temperatures fluctuations by days is shown in Figure 2. The number of hospitalizations by days is shown in Figure 3.

**Figure 1.**
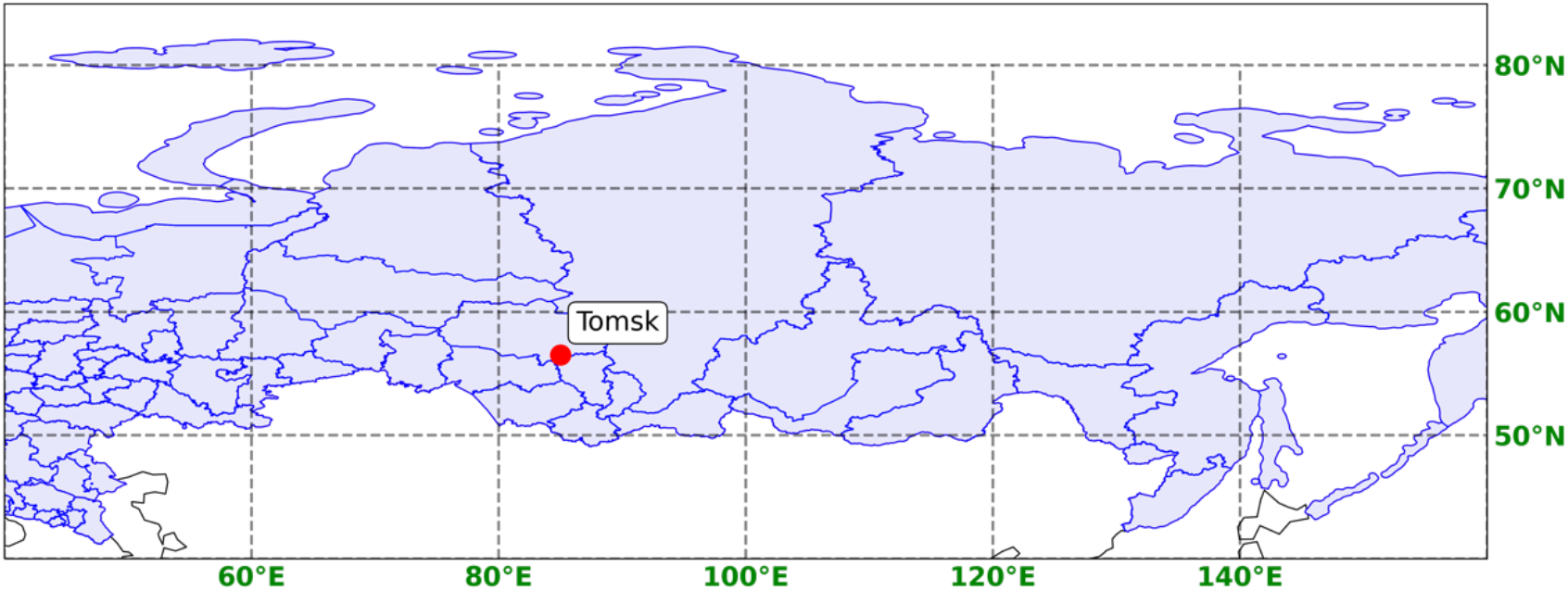
Map of Russia (Tomsk Oblast) The data of OpenStreetMap® was used, licensed under the Open Data Commons Open Database License (ODbL) by the OpenStreetMap Foundation (OSMF) and the Creative Commons Attribution-ShareAlike 2.0 license (CC BY-SA 2.0).

**Figure 2.**
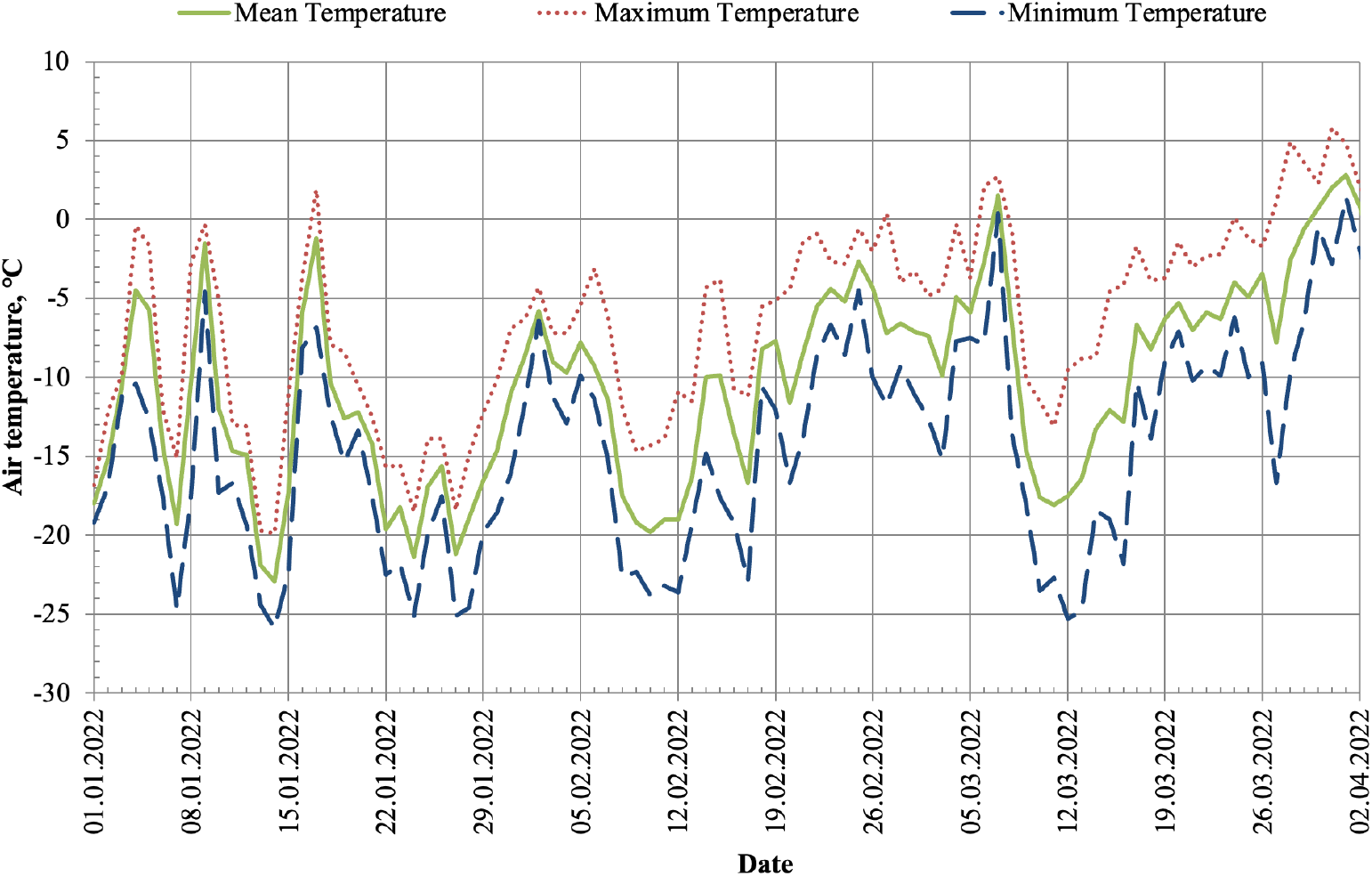
Outdoor daily temperature in Tomsk

**Figure 3.**
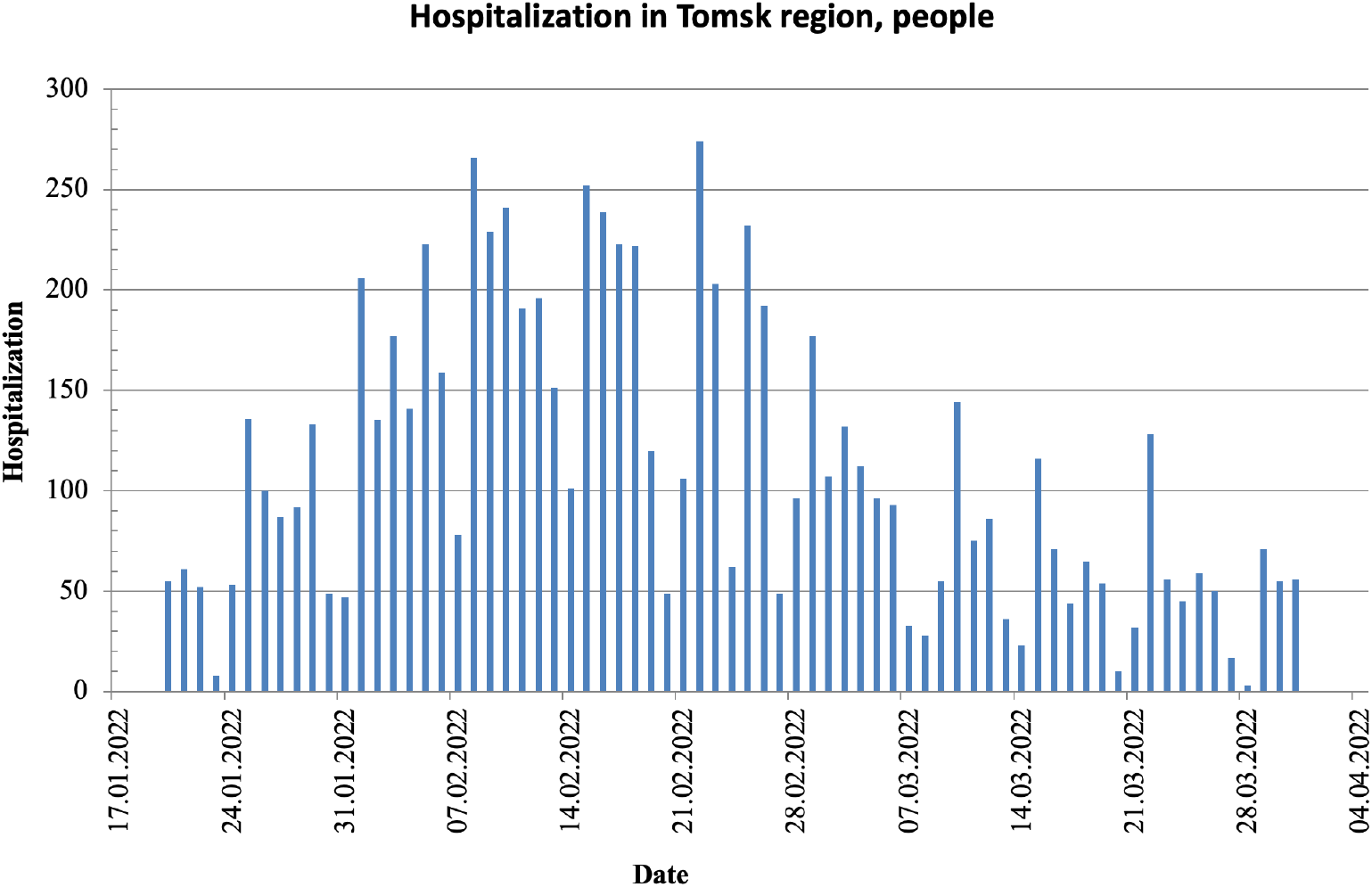
Daily hospitalizations in Tomsk

The wave of hospitalizations (Fig. 3) from January 20 (wave start) to March 31 (wave end) can be clearly associated with the main pandemic wave, and here we propose to consider this wave as a “main carrier wave” (see below), which depends on the emergence of a new strain and other global factors, including air temperature too.

From figure 2 it follows that the temperature was always low (below comfortable values for a person − this corresponds to the time of year for this region), which in itself is the main influencing factor. Therefore, here we consider only temperature fluctuations (temperature drop) as an additional factor that can make some adjustments to the health condition of infected individuals and, as a result, to “wave beats/bursts” that appear on this carrier wave (pandemic wave) in form of frequency modulation. **Conventionally, this approach can be compared with the principle of superposition of waves − when a “ripple”(frequency modulation) is superimposed on a carrier wave**.

As well as in wave dynamics, we initially believe that the potential of secondary bursts (modulation) has limited values (and we found confirmation of this, see below (table 1)). For example, if a surge is caused by some factor, then the subsequent factor may affect this surge too (the principle of superposition of waves), but this influence may not be so pronounced as the first factor influence was (the factor that acted as an initial trigger).

**Table 1.**
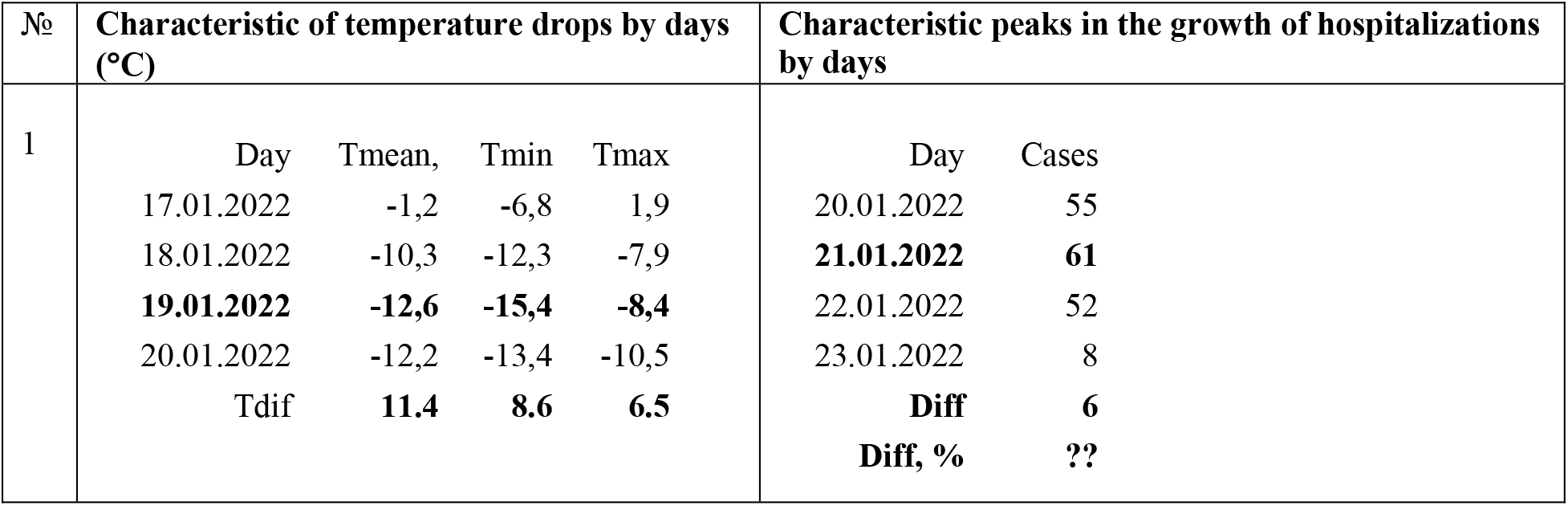
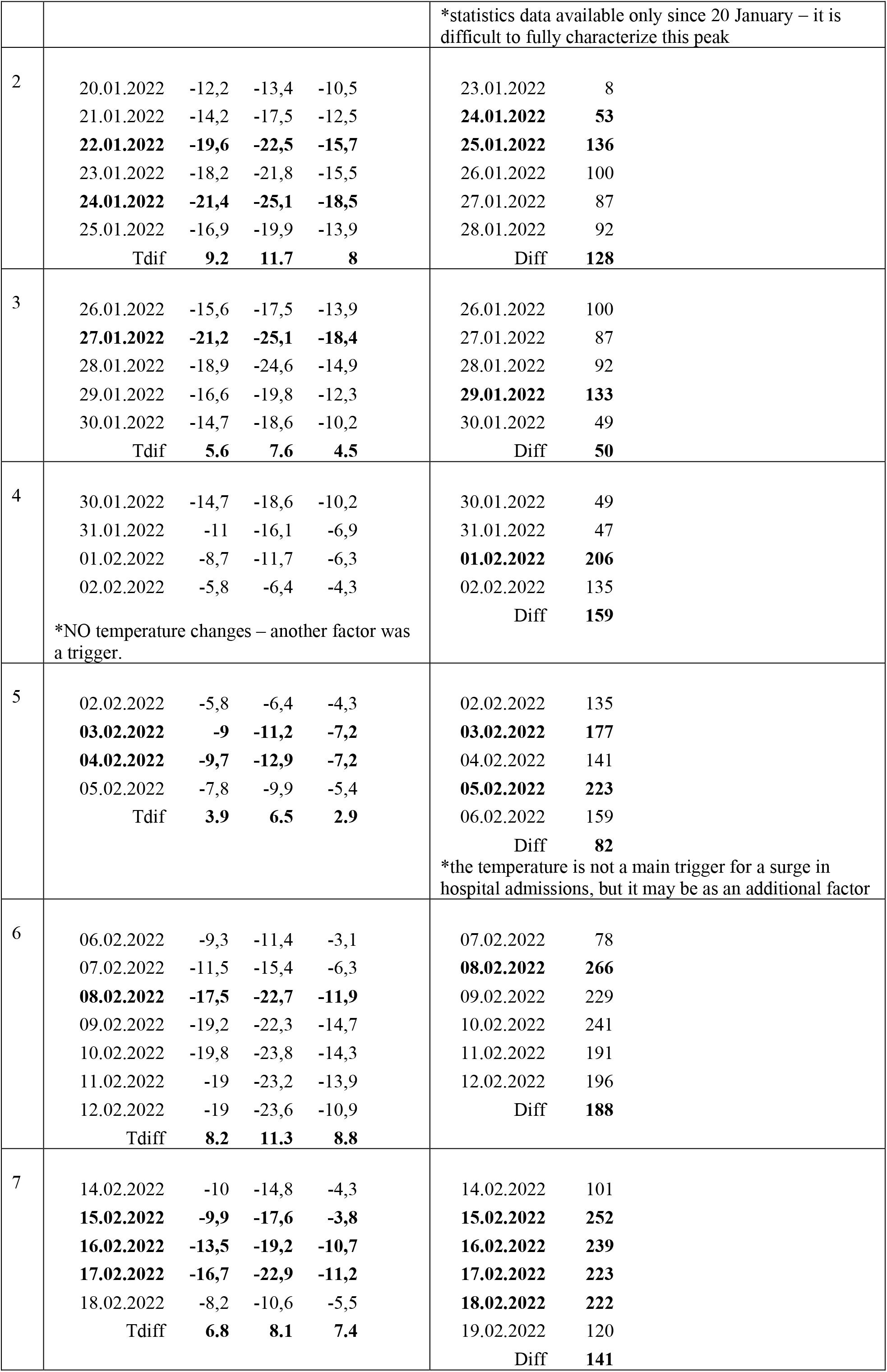
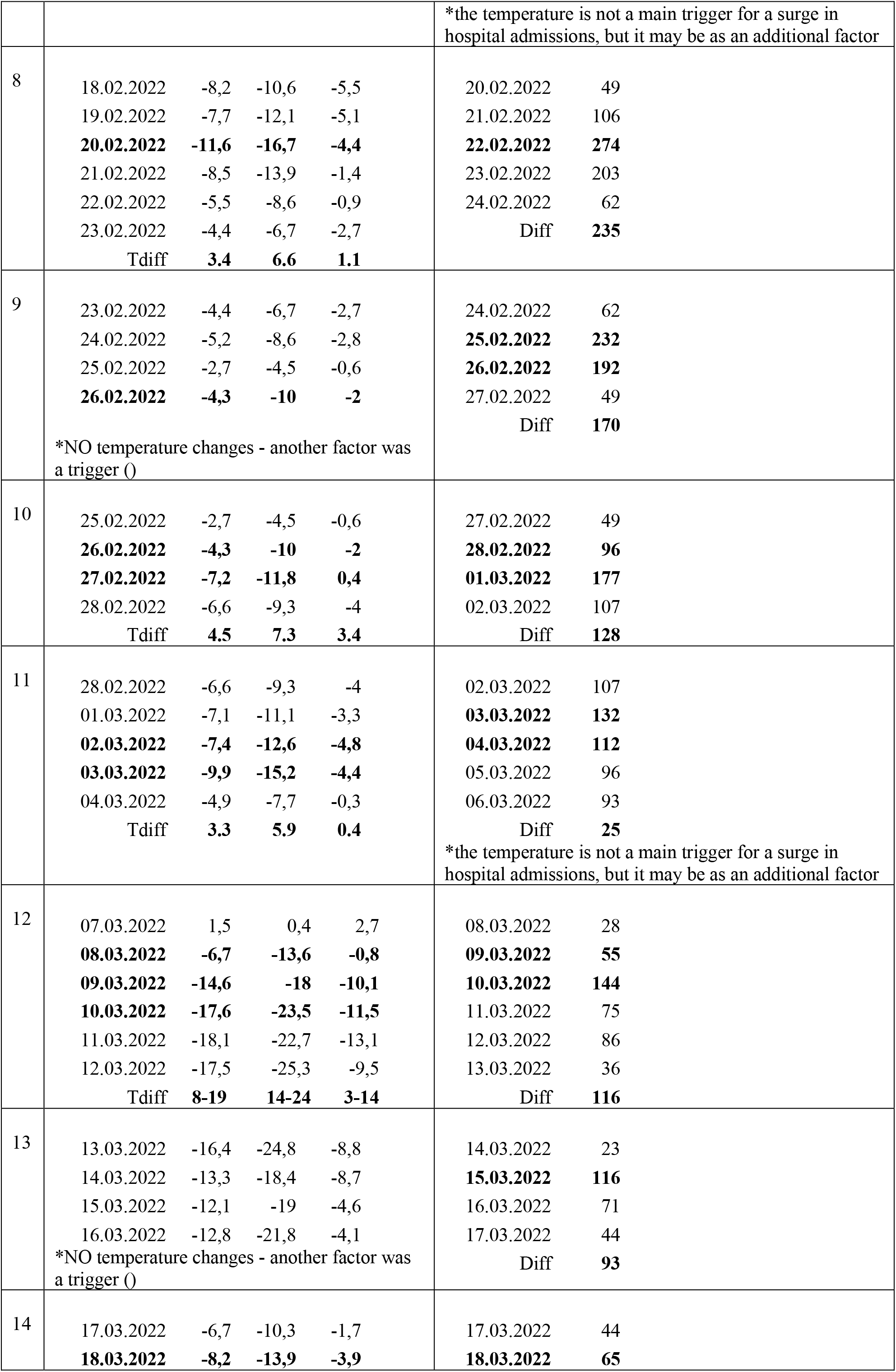
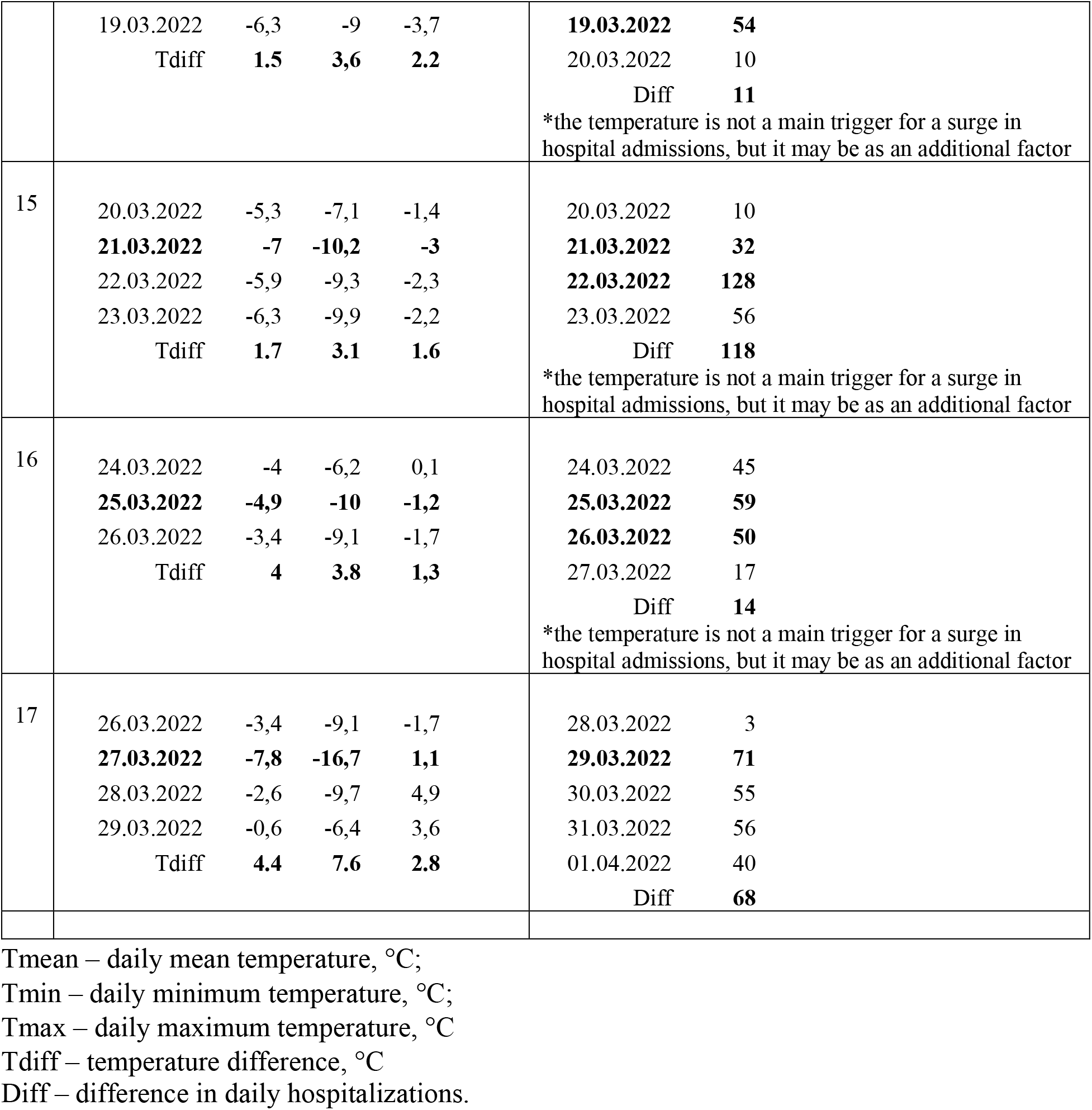
Characteristic points in fluctuations in ambient daily temperatures and daily hospitalizations.

In any case, many factors can act as triggers for wave modulation. Moreover, all the factors can act in a joint manner and overlap each other. For example, one factor (for example, temperature, or air humidity, or an air pollutant, or a weather parameter, etc.) triggers a surge in diseases/hospitalization wave, and then another factor appears and maintains this local surge on the main\ carrier wave.

At this stage, we have chosen the task of assessing the influence of air temperature as a simple and obvious acting factor. We believe that the effect of temperature should have an obvious and fast effect − and we found evidence for this in the results of the performed analysis (see below).

At the first stage, we looked for obvious and strong changes\drops in the daily temperature, and then compared them with the number of hospitalizations falling on those days, taking into account a possible time lag of several hours or days.

For the initial determination of the time lag, we chose the most characteristic points in the fig.2, where the temperature changed very strongly during one or two days, − that is, where such influence (drop in air temperature) is likely to be the most noticeable (even taking into account the other influencing factors). Since such a “cold” impact can and should affect the state of the “compromised” respiratory system (in accordance with the hypothesis put forward − see above).

In the next step, we compared changes in hospital admissions (fig. 3), which could correspond to the effects of temperature changes. Here, hospitalizations serve as an indicator of the severity of the disease and the deteriorating health of infected individuals. Thus, thanks to this preliminary stage, we found that the time lag is no more than 1-2 days.

Next, we carried out a detailed analysis of each change (drops) in ambient temperatures and analysis of each spike in the number of daily hospitalizations. The analysis is shown in table 1.

Thanks to the obtained analysis, we have identified 14 characteristic points where the temperature drop was more than 3 degrees (№ 1-3, 5-8, 10-17) (table 1)) – all of them were associated with characteristic bursts in the number of daily hospitalizations with a time lag of 1-2 days.

In 8 characteristic points of those 14 (№1-3, 6, 8, 10, 12, 13) the temperature can be characterized as a primary trigger (which acted as the main influencing factor).

In 6 characteristic points of those 14 (№5, 7, 11, 14-16) the temperature can be characterized as an additional factor (joint influence with other acting factors).

In total, 17 characteristic surges in the number of daily hospitalizations were identified over the entire period. Three of them (№3, 4, 9) had no connection with drop in daily temperature (the temperature did not change in these days) and were associated with other factors.

The data from the table once again confirmed the initially found value of the time lag of the short-term effect of temperature drops on worsening of disease in infected individuals (indicated by bursts of daily hospitalizations) − it averaged 1-2 days.

Thus, it was found that each characteristic points where the temperature drop was more than 3 degrees can be clearly associated with bursts in the number of daily hospitalizations.

It is important to note that since the pandemic started, many authors have analyzed the issue of impact of many factors (weather conditions, air pollution and etc.) on COVID-19 incidence, severity, and mortality, and literature linking to this issue has increased exponentially and now hundreds studies show the different results (see related revies in (Kang et al., 2021; Prévost et al., 2021; Liang & Yuan, 2022; Mandal et al., 2022; McClymont & Hu, 2021; Mejdoubi et al., 2020; Runkle et al., 2020; To et al., 2021; Tripathi et al., 2022; Xie & Zhu, 2020; Carballo et al., 2022; Culqui et al., 2022; Ishmatov, 2022; Kang et al., 2021; Perone, 2022; Taylor et al., 2022)). Most of the studies are statistical and analyze the correlation between various influencing factors. Moreover, many studies use limited datasets and are subject to several confounders, requiring caution when interpreting these results.

It is worth noting that today there are many studies of the correlation of weather factors and air temperature with COVID-19 in many countries and cities. Most of them agree that air temperature affects COVID-19 and authors describe some possible effects of air temperature on COVID-19 disease severity and transmission rates (see related reviews in (Kang et al., 2021; Prévost et al., 2021; Liang & Yuan, 2022; Mandal et al., 2022; McClymont & Hu, 2021; Mejdoubi et al., 2020; Runkle et al., 2020; Tripathi et al., 2022).

Most of the studies are statistical and have large-scale character – entire countries are studied, many variables are described simultaneously, and long periods are examined. Our study is based on a mechanistic approach − we did not just choose a parameter for correlation analysis, but proposed a clear and precise mechanism of how this parameter can affect and what this effect can lead to. Our study can be characterized by strong concretization of the chosen direction of research, − limited to one city (where the population is evenly exposed to the same impacts (conditionally) and one specific pandemic wave (when one strain of the virus prevails)). Perhaps this approach made it possible to discover the relationships and patterns in this study, which, perhaps, would not be so pronounced for large-scale study.

### Future directions and limitations

Obviously, not every external influence can have the same pronounced effect as the effect of the air temperature. Sunlight, precipitation, humidity, wind speed and etc. − these are indicators that are difficult to consider and analyze, since it is now difficult to associate them with effects on the respiratory system, within the framework of a mechanistic approach proposed in this study.

Here it is important to consider the air humidity (as a main weather parameter). We examined this indicator in detail and came to the conclusion that it is difficult task for a separate analysis and a detailed study of this issue are required.

On the one hand the absolute humidity (AH) is directly related to air temperature. Relative humidity (RH) is not so clearly related to temperature, but with sudden changes in daily temperature, this relationship is direct (especially for continental climates). For example, the maximum moisture content possible in air varies with temperature and as a result, the variation of ambient temperature can significantly affects the ambient RH.

On the other hand, air humidity should affect the efficiency of heat and mass transfer in the respiratory tract – thereby affecting the cooling efficiency of the airways epithelial cells. Moreover, low RH should affect the effectiveness of mucosal drying and, as a consequence, affect the mucociliary clearance.

But in any case, here relative humidity should always be considered together with temperature – since temperature (as well as humidity) plays a significant role in heat and mass transfer processes. This aspect (within the impact on COVID-19) need to be investigated in the future studies.

It is important to note that the conception of the effect of “supersaturation in the airways” which was proposed in (Ishmatov, 2019; Ishmatov, 2020b) is most promising here − because it links temperature and RH into one mathematical function. The main sense of the supersaturation in the airways is that this effect depends simultaneously on both temperature and RH of inhaled air. Thus, temperature and RH are the parameters of one simple function (it is the effect of supersaturation) and no longer need to consider the separate influence of humidity or temperature on the human health.

Cold, rainy or wet weather can induce supersaturated conditions in the airways. It was shown that supersaturation can lead to enhanced deposition of inhaled ambient aerosols, infectious aerosols and air pollutants in the human airways (Ishmatov, 2017a; Ishmatov, 2020b). Xu et al. (2021) also confirmed this aspect in the study with the help of numerical experiments.

Moreover, role of effect of supersaturation in a critical cooling of local areas in the human upper airways was discussed, and this involves cold stress and reducing, as a consequence, the antiviral immune response of the airway epithelial cells. The additional acidification of epithelial lining fluid in the local areas of the respiratory tract was also considered. Such acidic stress may lead to destructive impacts on the respiratory cells and weakening of the defense mechanisms of the airways (Ishmatov, 2017b; Ishmatov, 2020b).

According to the above, the supersaturation effect may be used in the future for predictions and analysis of the changing epidemiological situation (the first attempt was made in (Ishmatov, 2017c)). However, these aspects are preliminary and need to be investigated in the future studies.

In our opinion, of all potential influencing factors, air pollution fits the mechanistic approach proposed in this study. Air pollution here suggests the same effect as temperature since it directly affects the human respiratory system. Research is currently underway in this direction and first preliminary results was presented in (Ishmatov et al., 2022).

## Conclusion

Many factors influence the pathogenesis and epidemiology of COVID-19. Considering the whole set of influencing factors at the same time can be quite difficult and it is counterproductive now and therefore it is important to identify the all factors and understand how they mechanistically influence COVID-19 and to evaluate the extent of this impact. It is a rigorous element-by-element analysis of each influencing factor that can now give good results. In the end, thanks to a mechanistic approach, in the future we will be able to assemble the whole epidemiological puzzle brick by brick.

We believe that it was a good idea to conduct research using the hypothesis of the ‘compromised airway epithelium’. The ‘compromised airway epithelium’ of infected persons can be extremely sensitive to external influences and therefore can serve as a good indicator of the impact on the respiratory system of external factors. And the time lag of the impacts is rather short (1-2 days), which gives ample opportunities for analyzing and predicting the consequences of such impacts.

Also, the idea of using the wave theory, the principle of superpositions and frequency modulation, together with the mechanistic approach, looks quite promising in the analysis of epidemic waves. But this requires a strictly mathematical approach to the analysis of all influencing factors in the future studies.

In this study we have chosen the task of assessing the influence of air temperature on the respiratory system as a simple and obvious acting factor. It was found that even a small and/or short-term impact of drop in outdoor daily temperatures on the ‘compromised airway epithelium’ lead to increased symptoms and severity of the disease (COVID-19). Changes in the ambient daily temperature are well predicted for days and weeks by modern methods. And this aspect can be used to improve systems to prevent additional risks connected with impact of drop in air temperature on worsening disease in patients and infected people who do not have or have mild or subtle symptoms of the disease – especially during an epidemic or pandemic wave. It was found that the results of the study can be used in predicting a sudden increase in the number of hospitalizations, which can be used to timely warn clinics and medical hospitals for an increase in the number of seriously ill patients. Thus the findings are another brick in the wall and besides the above, they can also be used in preparing health care systems and populations to better protect those most at risk from serious complications from becoming infected, and support timing of interventions to help achieve this.

## Data Availability

All data produced in the present work are contained in the manuscript

## Acknowledgments

We thank the “CityAir” and Pavel Glotov (General director of “CityAir”) for providing measured meteorological data from weather station located in Tomsk.

***The study was supported by a grant of the Russian Science Foundation No 22-27-20111, https://rscf.ru/project/22-27-20111/ and funded by the Administration of the Tomsk Region***.

